# BNT162b2 COVID-19 mRNA vaccine elicits a rapid and synchronized antibody response in blood and milk of breastfeeding women

**DOI:** 10.1101/2021.03.06.21252603

**Authors:** Michal Rosenberg Friedman, Aya Kigel, Yael Bahar, Yariv Yogev, Yael Dror, Ronit Lubetzky, Ariel Many, Yariv Wine

## Abstract

We describe the dynamics of the vaccine-specific antibody response in the breastmilk and serum in a prospective cohort of ten lactating women who received two doses of the Pfizer-BioNTech BNT162b2 COVID-19 mRNA vaccine. The antibody response was rapid and highly synchronized between breastmilk and serum, reaching stabilization 14 days after the second dose. The predominant serum antibody was IgG. The response in the breastmilk included both IgG and IgA with neutralizing capacity.

---

The accelerated COVID-19 vaccination campaign in Israel was initiated in December 2020 and currently nearly 50% of the adult population have received the Pfizer-BioNTech BNT162b2 COVID-19 mRNA vaccine (mRNA vaccine)^1^. The vaccine campaign initially targeted high-risk populations (≥60 years old and healthcare providers)^2^ and was soon expanded to lactating women^3^. Notwithstanding the reported high efficiency of this vaccine^4^ and evidence for the generation of viral-specific antibodies in the breastmilk of women with COVID-19^5-7^, there are no available data on its efficiency in lactating women or its potential benefits in neonatal protection via the passive transfer of vaccine-specific antibodies in breastmilk^8^. The current knowledge gap is preventing global health authorities from making concrete recommendations regarding vaccination during lactation.

We describe the dynamics of the breastmilk and serum antibody response in a prospective cohort of ten lactating healthcare providers, mean age 34.6 (range 30-38), who received the first dose of the mRNA vaccine approximately five months postpartum (mean 154 days, range 68-382) and the second dose 21 days later (**Supplementary Table 1)**.

To obtain calculated endpoint titers for IgG and IgA in breastmilk and serum dyads against the SARS-CoV-2 spike and receptor binding domain (RBD) proteins, serial dilution ELISAs were run on days 7 and 14 after the first (designated 1D7 and 1D14, respectively) and second (designated 2D7 and 2D14, respectively) vaccine doses (Supplementary Fig. 1 and Supplementary Fig. 2). We found that the spike (**Fig. 1**) and RBD-specific (**Supplementary Fig. 3**) antibody responses in breastmilk and serum were synchronized for IgG and IgA.

**Fig. 1.**
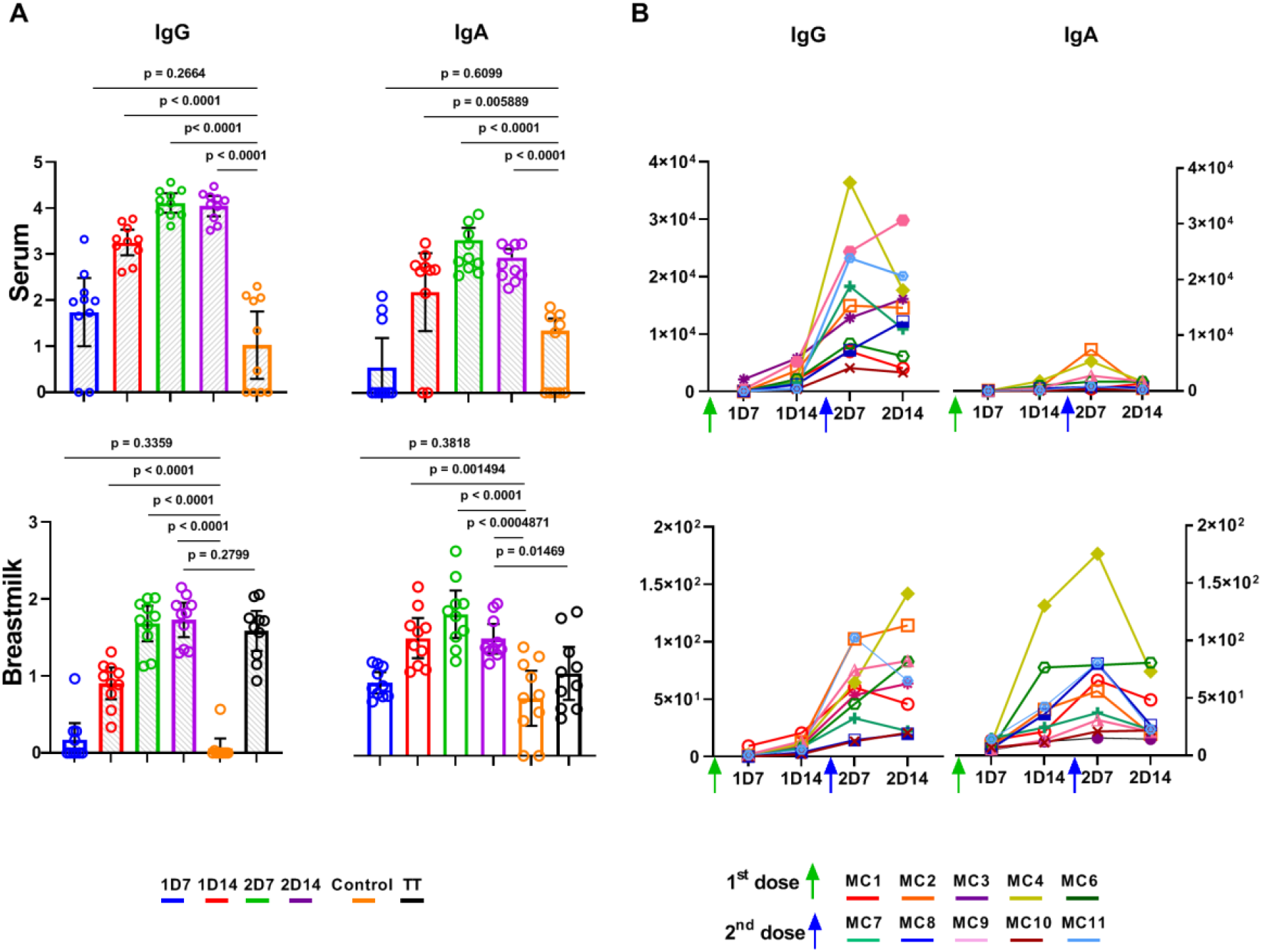
Endpoint titers of spike-specific IgG and IgA antibodies in breastmilk and serum of mothers vaccinated with the Pfizer-BioNTech vaccine (n=10). (**A**) Endpoint titers were interpolated by applying a four-parameter logistic curve on reciprocal dilution series for IgG and IgA for all serum and lactoserum samples at four time points (by color) and for tetanus toxoid (TT)-specific IgG and IgA in breastmilk. P values were determined with an unpaired, two-sided Mann-Whitney U-test after applying Bonferroni correction; P < 0.0083 was considered statistically significant. Results are presented as geometric means and 95% confidence intervals. Y-axis units are endpoint titers on a logarithmic scale. Control serum (n=10) and lactoserum samples (n=10) were obtained before the COVID-19 pandemic. (**B**) Endpoint titers per participant, each shown in a different color. Green and blue arrows indicate the time points of administration of the first (t=0) and second (t=21) vaccine doses, respectively. Y-axis units are endpoint titers on a linear scale.

At 1D7, the vaccine-specific endpoint titers had not increased significantly above the titers in the control group (pre-pandemic serum and lactoserum), and the first significant increase in antibody titers was evident at 1D14. The upward trend peaked at 2D7, followed by a slight decrease in titers at 2D14.

Since all participants had received the tetanus, diphtheria, acellular pertussis (Tdap) vaccine during the third trimester^9^, we were able to compare the breastmilk endpoint titers for IgG and IgA elicited by the mRNA vaccine at 2D14 to tetanus toxoid (TT)-specific antibody titers in the same participants. We found that the spike- and RBD-specific IgG and IgA titers did not differ significantly from the TT-specific antibody titers.

Thereafter, we evaluated the vaccine-specific IgG/IgA ratio in breastmilk and serum. The serum antibody response was dominated by IgG, and the IgG/IgA ratio was significantly higher in serum than in breastmilk at all four time points (**Supplementary Fig. 4**). The IgG/IgA ratio in breastmilk indicated that the vaccine-specific response was not dominated by IgA, but rather that the IgG/IgA ratio increased over time, as previously described following respiratory syncytial virus immunization^10^.

To better understand the temporal dynamics of the antibody response following mRNA vaccination, we calculated the fold-change of endpoint titers at each time point compared to the preceding time point. The vaccine-specific IgG and IgA titers in breastmilk and serum increased substantially 7 days following each vaccine dose, and the increase in fold-change halted 14 days following the second dose. Noteworthy, for spike- and RBD-specific IgA in breastmilk, the fold-change at 2D14 fell below 1, indicating declining IgA levels at that time point (**Supplementary Fig. 5**). Calculation of fold-change above the endpoint titer at 1D7 (**Fig. 2, Supplementary Fig.6**) showed that the dynamics of the vaccine-specific response in both serum and breastmilk stabilized at 2D14, since the fold-change at 2D14 was not significantly higher than that at the preceding time point.

**Fig. 2.**
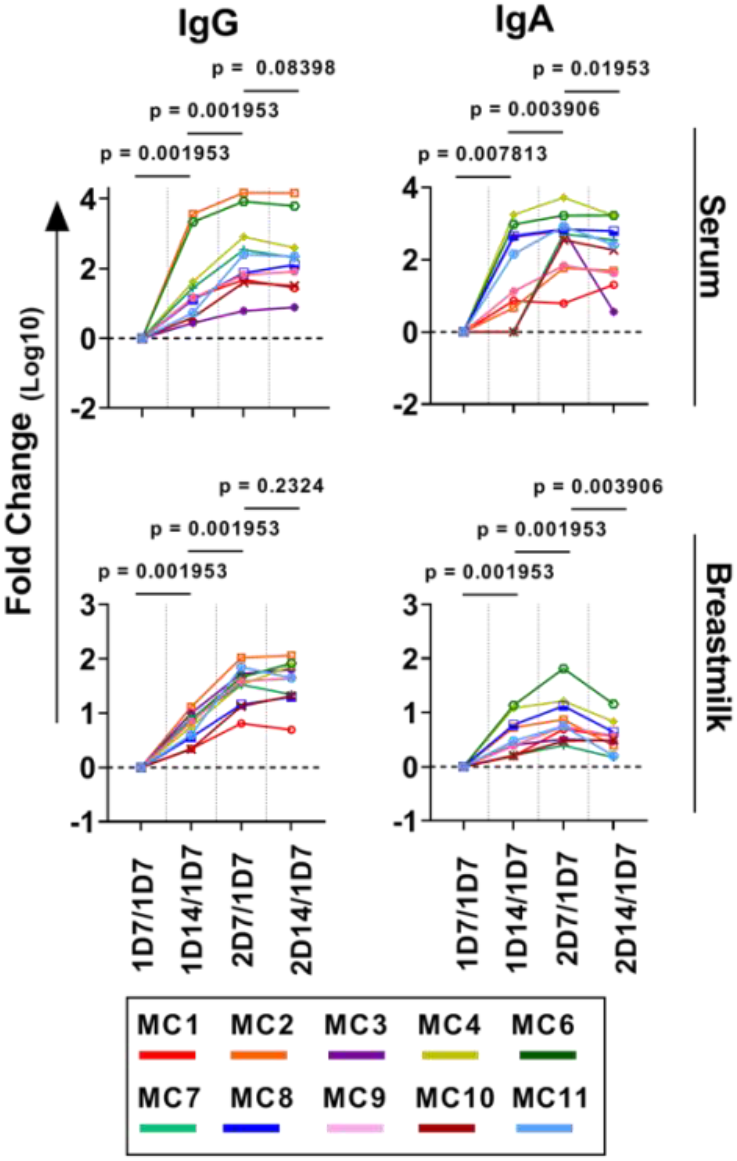
Dynamics of the vaccine-specific antibody response and the antibody neutralization capacity. Fold-changes in antibody titers compared to the first time point are plotted by patient. Comparisons between the fold-change in vaccine-specific IgG and IgA antibodies were made using the Wilcoxon signed-rank test; P < 0.05 was considered statistically significant. Y-axis units are fold-change on a logarithmic scale.

Finally, we evaluated the fraction of neutralizing antibodies in breastmilk with a previously developed in vitro competitive-blocking assay^11^. Both anti-spike IgG and IgA in the breastmilk of all participants exhibited a potential neutralization capacity [on average, 13% of the vaccine-specific antibodies were blocked in the assay (**Supplementary Fig. 7**]. Of note, a recent publication^6^ reported that only 62% of breastmilk samples (containing antibodies) from women with COVID-19 were found to have neutralizing capacity in vitro.

The importance of breastfeeding in early infancy is highlighted by the strong correlation between breastfeeding and the overwhelming decline in risks of infection and infection-associated morbidity and mortality^12,13^: Breastfeeding has been associated with a decrease in the number of cases of respiratory illness^14^, a decreased risk of hospitalization for respiratory diseases^15,16^, and protection against a wide range of infections that may colonize the gut^17,18^. Reported cases of COVID-19 patients who experienced GI -related symptoms^19^ and the detection of viral RNA in stool specimens^20^ suggest a possible fecal-oral transmission route^21^.

In summary, the study provides evidence for the rapid production of vaccine-specific antibodies, both IgA and IgG. Moreover, neutralizing capacity was observed in all samples. This study also indicates the potential protection of breastfed infants by administration of the BNT162b2 COVID-19 vaccine to the breastfeeding mother.

## Data Availability

Data are available on request due to privacy or other restrictions

## Acknowledgements

This work was supported by the Israeli Ministry of Health (MOH) grant #3-17162)

## Contributions

The study was initiated by M.R.F., A.M. and Y.W. Sample collection and processing was carried out by M.R.F., A.K., Y.D. Experiments were carried out by A.K., Y.B. and Y.D. The original manuscript draft was written by M.R.F., A.K., A.M. and Y.W. Review and editing of the manuscript were carried out by M.R.F., A.K., Y.B., Y.D., R.L., Y.Y., A.M. and Y.W.

## Notes

### Competing Interest Statement

The authors have declared no competing interest.

### Funding Statement

This work was supported by the Israeli Ministry of Health (MOH) grant #3-0000-17162)

### Author Declarations

Sample collection was performed under the following ethical approvals: 1/ Institutional review board (IRB) approvals number 0002269-4 and 0002757-1 given at Tel Aviv University. Chair of IRB committee Prof. Meir Lahav, Faculty Medicine, Tel Aviv University. 2/ Ethical approval number 1088-20-TLV given at Tel Aviv Sourasky medical center. Prof. Shmuel Kivity, Chairman, Institutional Review Board (IRB) / Ethics (Helsinki) Committee,Tel Aviv Sourasky medical center and under approval by the director of Tel Aviv Sourasky medical center, Prof. Ronni Gamzu. All participants provided informed consent for the use of their data and clinical samples for the purposes of the present study. Study participant's data includes less than three indirect identifiers throughout the manuscript in compliance with ethical guidelines.

