## Supplemental file for "BNT162b2 COVID-19 mRNA vaccine elicits a rapid and synchronized antibody response in blood and milk of breastfeeding women"

Supplementary

**Human subjects, sample collection and processing**

The cohort comprised of ten lactating healthcare providers received two doses of the Pfizer-BioNTech BNT162b2 COVID-19 mRNA vaccine (mRNA vaccine) twice at a 21-days interval between the first and second doses. All participants provided informed consent for the use of their data and clinical samples for the purposes of the present study. Sample collection was performed under institutional review board approvals number 0002269-4 and 0002757-1 given at Tel Aviv University and under ethical approval number 1088-20-TLV given at Tel Aviv Sourasky medical center. Breastmilk and blood dyads were collected from the COVID-19 vaccinees into BD vacutainer K2 EDTA collection tubes and sterile containers, respectively. Sample dyads were collected at four time points, namely, 7 and 14 days following the first and second vaccine doses designated as 1D7, 1D14, 2D7 and 2D14, respectively. Control serum and lactoserum samples were obtained from 10 healthy individuals collected prior the COVID-19 pandemic.

Isolation of plasma from whole blood was performed by density gradient centrifugation, using Uni-SepMAXI$^+$ lymphocyte separation tubes (Novamed) according to the manufacturer's protocol. Breastmilk lactoserum was separated from whole milk by centrifugation in 50-mL conical tubes at 500 $\times$ g, swinging bucket, Room Temperature (RT), 20 min, acceleration = 9, brake = 1. The upper lipid layer was discarded, and the lactoserum was transferred to a clean 50-ml tube. All serum and lactoserum samples were stored

at -20°C.

**Expression and purification of recombinant protein**. The plasmids for expression of recombinant SARS-CoV-2 receptor-binding domain (RBD) and spike protein were kindly provided by Dr. Florian Krammer, Department of Microbiology, Icahn School of Medicine at Mount Sinai, New York, NY, USA. The RBD sequence is based on the genomic sequence of the first virus isolate, Wuhan-Hu-1, which was released on 10 January 2020[1]. The plasmids for the expression of recombinant human angiotensin I converting enzyme 2 (hACE2) was kindly provided by Dr. Ronit Rosenfeld, Israel Institute for Biological Research (IIBR). The cloned region encodes amino acids 1-740 of hACE2 followed by 8xHis-tag and a Strep Tag at the 3' end, cloned in a pCDNA3.1 backbone. Recombinant RBD and hACE2 were produced in Expi293F cells (ThermoFisher Scientific) by transfection of these cells with a purified mammalian expression vector using an ExpiFectamine 293 Transfection Kit (Thermo Fisher Scientific), according to the manufacturer's protocol, and as described previously[1]. Supernatants from transfected cells were purified on a HisTrap affinity column (GE Healthcare) using a 2-step elution protocol with 5 column volumes (CV) of elution buffer supplemented with 50mM imidazole in phosphate-buffered saline (PBS), pH 7.4 followed by 250mM imidazole in PBS, pH 7.4, for RBD and spike protein and for hACE2 by 500 mM imidazole in PBS. Elution fractions containing clean recombinant proteins were merged and dialyzed using Amicon Ultra (Mercury) cutoff 10K against PBS (pH 7.4). Dialysis products were analyzed by 12% SDS–PAGE for purity, and concentration was measured using Take-5 (BioTek Instruments).

**Serum titer measurement and *in vitro* competition assay.** Serum IgG and IgA antibody endpoint titers were measured by enzyme-linked immunosorbent assay (ELISA). Spike$^+$ and RBD$^+$ Ig in lactoserum and serum were determined using half-area 96-well ELISA plates (Greiner Bio-One) that had been coated overnight at 4 °C with 2 µg/ml RBD or spike proteins in PBS (pH 7.4). Thereafter, the coating solution was discarded, and the ELISA plates were blocked with 150 µl of 3% w/v skim milk in PBS for 1 h at 37 °C. After discarding the blocking solution, duplicates of 36 µl of serum diluted 1:100 or 36 µl of lactoserum diluted 1:1 in 3% w/v skim milk in PBS were added to the first row of the coated plate. Dilutions were carried out with a threefold dilution factor, and the plates were incubated for 1 h at RT. Then, plates were washed three times with 0.05% PBS-Tween 20 (PBST) and incubated for 1 h at RT with horseradish peroxidase (HRP) conjugated anti-human IgG (Jackson Immunoresearch, #CAT 109035003) /anti-human IgA (Jackson Immunoresearch, #CAT 109035011) (25 µl, 1:5000 ratio in 3% w/v skim milk in PBS). Plates were then subjected to three washing cycles with 0.05% PBST, and developing was carried out by adding 25 µl of 3,3',5,5'-tetramethylbenzidine (TMB), followed by quenching with 25 µl of 1 M sulfuric acid. Plates were read using the Epoch Microplate Spectrophotometer ELISA plate reader at a wavelength of 450 nm.

The competitive ELISA for lactoserum was carried out using half-area 96-well ELISA plates that had been coated overnight at 4 °C with 25 µl of 2 µg/ml spike protein in PBS (pH 7.4). The following day, the coating solution was discarded, and ELISA plates were blocked with 150 µl of 3% w/v skim milk in PBS for 1 h at 37 °C. The blocking solution was discarded, 25 µl of 300 nM hACE2 in 3% w/v skim milk were added to the positive hACE2 wells, and 3% w/v skim milk in PBS was added to the negative hACE2 wells for 1 h at RT. Thereafter, duplicates of 1:1 diluted lactoserum samples with and without 300 nM hACE2 were added to the positive/negative hACE2 wells (respectively) and serially diluted threefold in 3% w/v skim milk in PBS. Plates were incubated for 1 h at RT and then washed three times with PBST. For the detection phase, 25 µl of HRP conjugated anti-human IgG or anti-human IgA secondary antibody were added (1:5000 ratio in 3% w/v skim milk in PBS), and plates were incubated for 1 h at RT, followed by three washing cycles with 0.05% PBST. Developing was carried out by adding 25 µl of TMB, and the reaction was quenched by adding 25 µl of 1 M sulfuric acid. Plates were read using the Epoch Microplate Spectrophotometer ELISA plate reader at a wavelength of 450 nm.

Endpoint titers were determined as the maximum serum/lactoserum dilution with an $O.D._{450}$ signal that is 3 standard deviations above background. The Mann-Whitney test was used to compare continuous variables of two independent groups, and significance was set at P = 0.0083 following Bonferroni correction. The Wilcoxon signed-rank test was used to compare matched samples. All reported P values were two-tailed. All statistics were performed with GraphPad Prism 9.0.2 (GraphPad Software).

Supplementary Table 1 | Study cohort information. Mean age 34.6 (range 30-38). Days from birth to first dose of BNT162b2 mRNA vaccine - mean 154 days (range 68-382)

| Patient | Gravidity Parity (GP) | Gesta-tional age at delivery | From birth to first dose of BNT162b2 mRNA vaccine (days ) | Infant nutrition | From TdaP vaccine to day 7 (months) | From last COVID-19 RT-PCR to first dose of vaccine (weeks) |
|---|---|---|---|---|---|---|
| MC1 | G3P3 | 39+5 | 382 | MOM, solid food | 13 | 6 |
| MC2 | G5P3 | 39+0 | 109 | MOM | 6 | 2 |
| MC3 | G1P1 | 38+4 | 164 | MOM, solid food | 7 | 2 |
| MC4 | G3P2 | 37+6 | 68 | MOM + formula | 4 | 10 |
| MC6 | G2P1 | 40+6 | 152 | MOM + formula | 6 | 14 |
| MC7 | G4P3 | 40+0 | 148 | MOM, solid food | 8 | 21 |
| MC8 | G4P2 | 38+5 | 81 | MOM | 4 | 11 |
| MC9 | G2P2 | 40+0 | 143 | MOM | 6 | 0.5 |
| MC10 | G3P3 | 40+0 | 135 | MOM, solid food | 6 | 19 |
| MC11 | G2P2 | 38+4 | 159 | MOM, solid food | 7 | 23 |

* No infant co-morbidities.

MOM – mother's own milk.

Supplementary Fig. 1

A

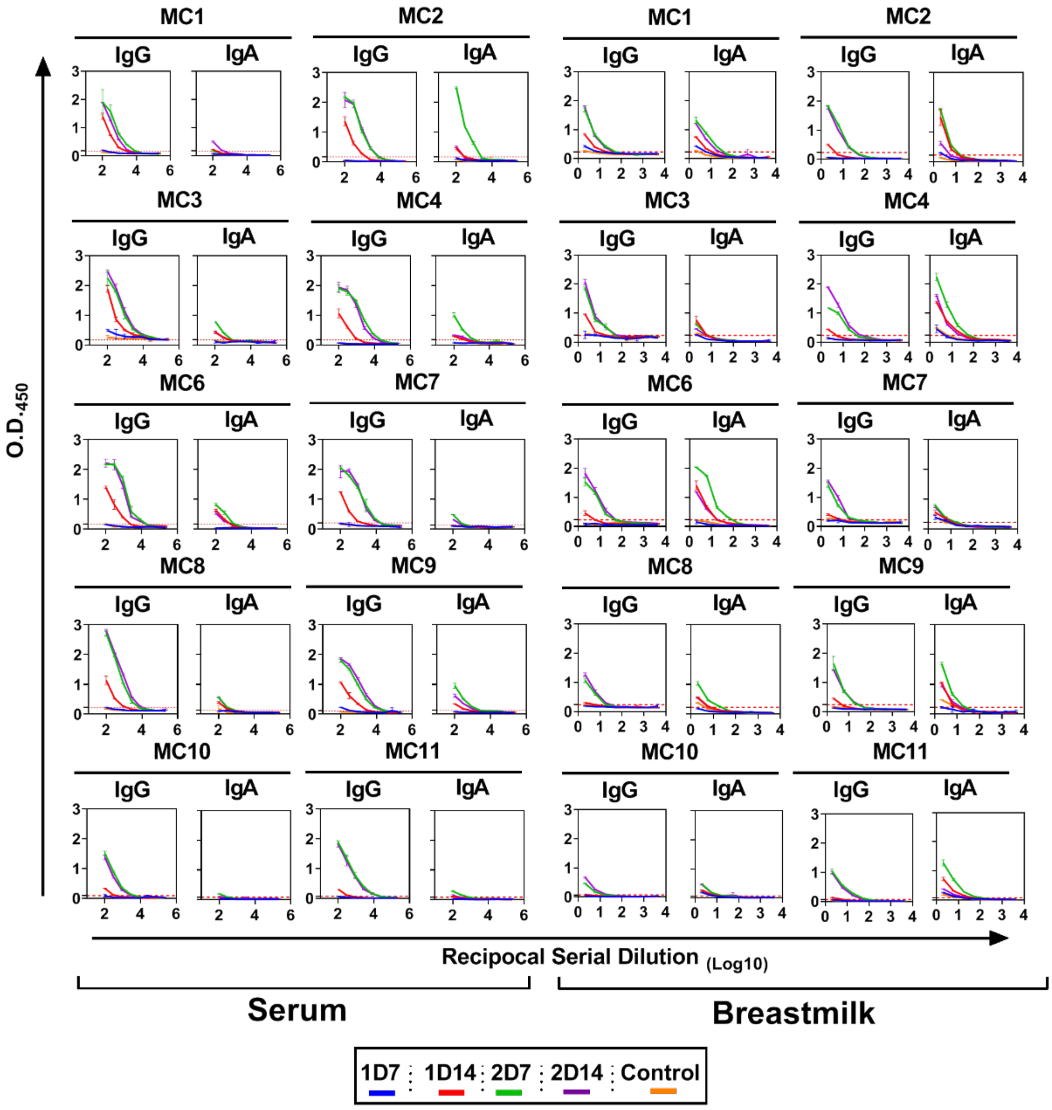

B

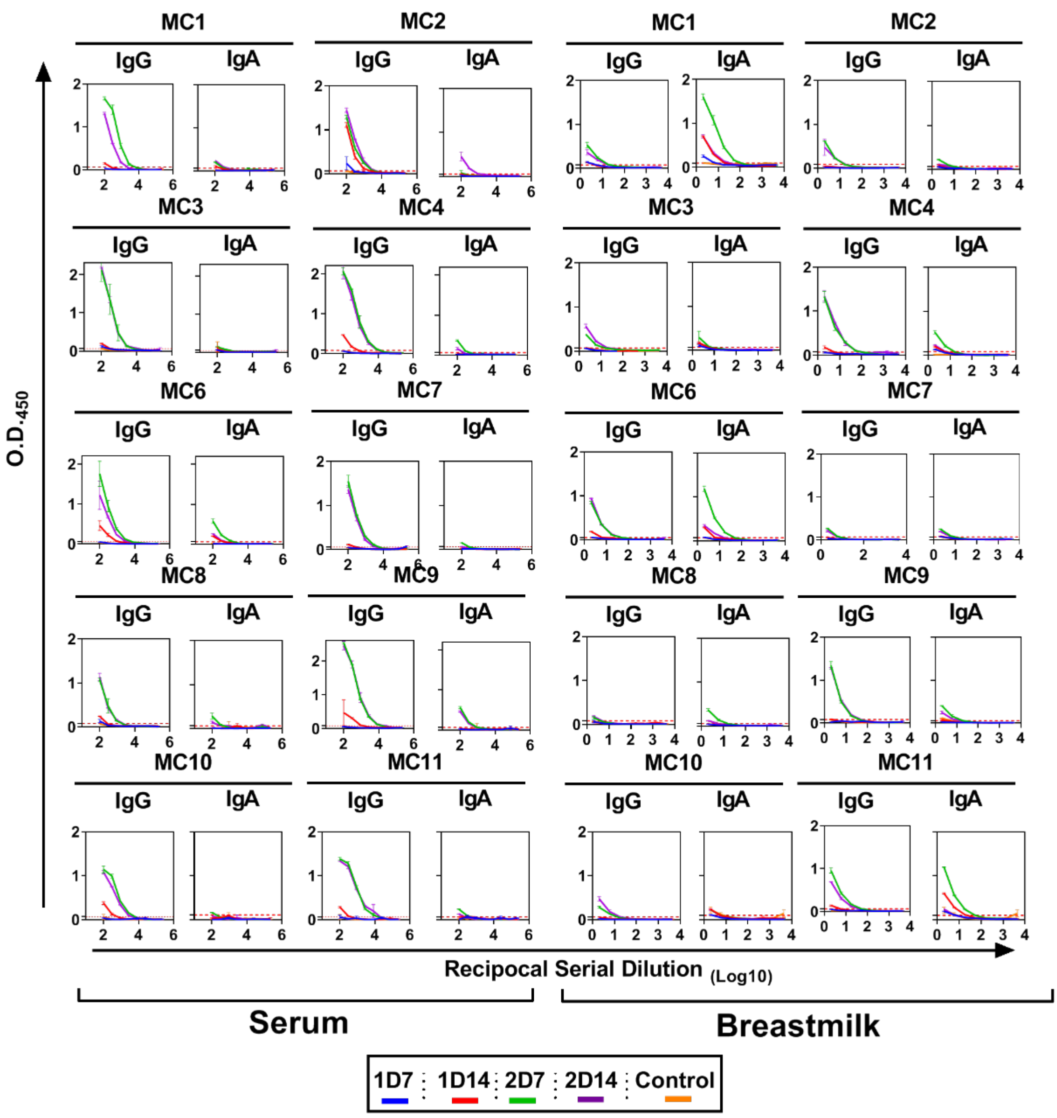


**Supplementary Fig. 1 | Serial dilution ELISA to determine endpoint titers.** Each plotted graph includes obtained O.D.$_{450}$ values that were subtracted from blank values for each plate. ELISA graphs include serum or breastmilk samples obtained at 4 time points (by color) for each vaccinee and a pre-pandemic negative control that were tested against the SARS-CoV-2. (**A**) spike protein and (**B**) RBD. Dashed red lines represent the threshold value equivalent to background + 3 s.d. values.

Supplementary Fig. 2

A

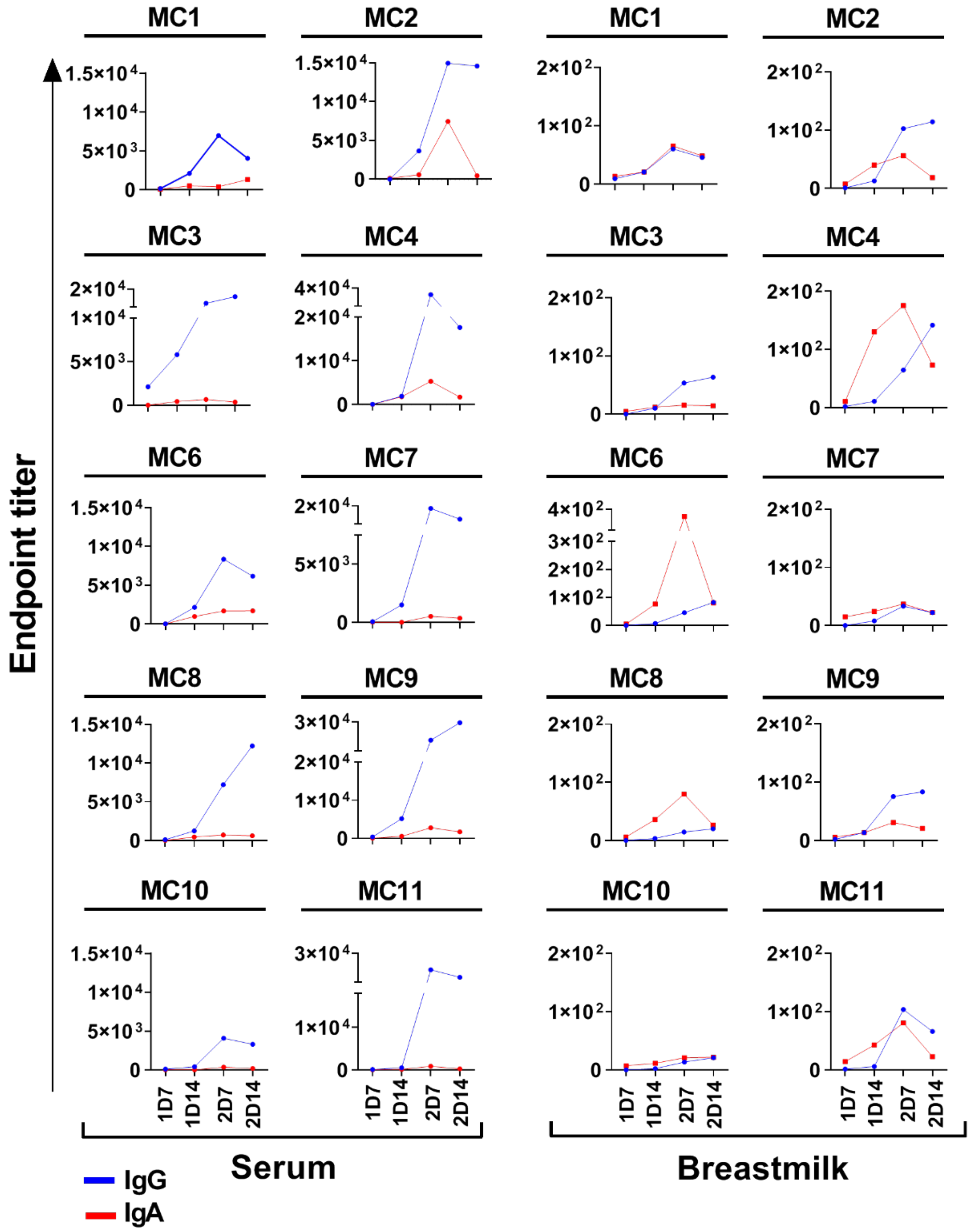

**B**

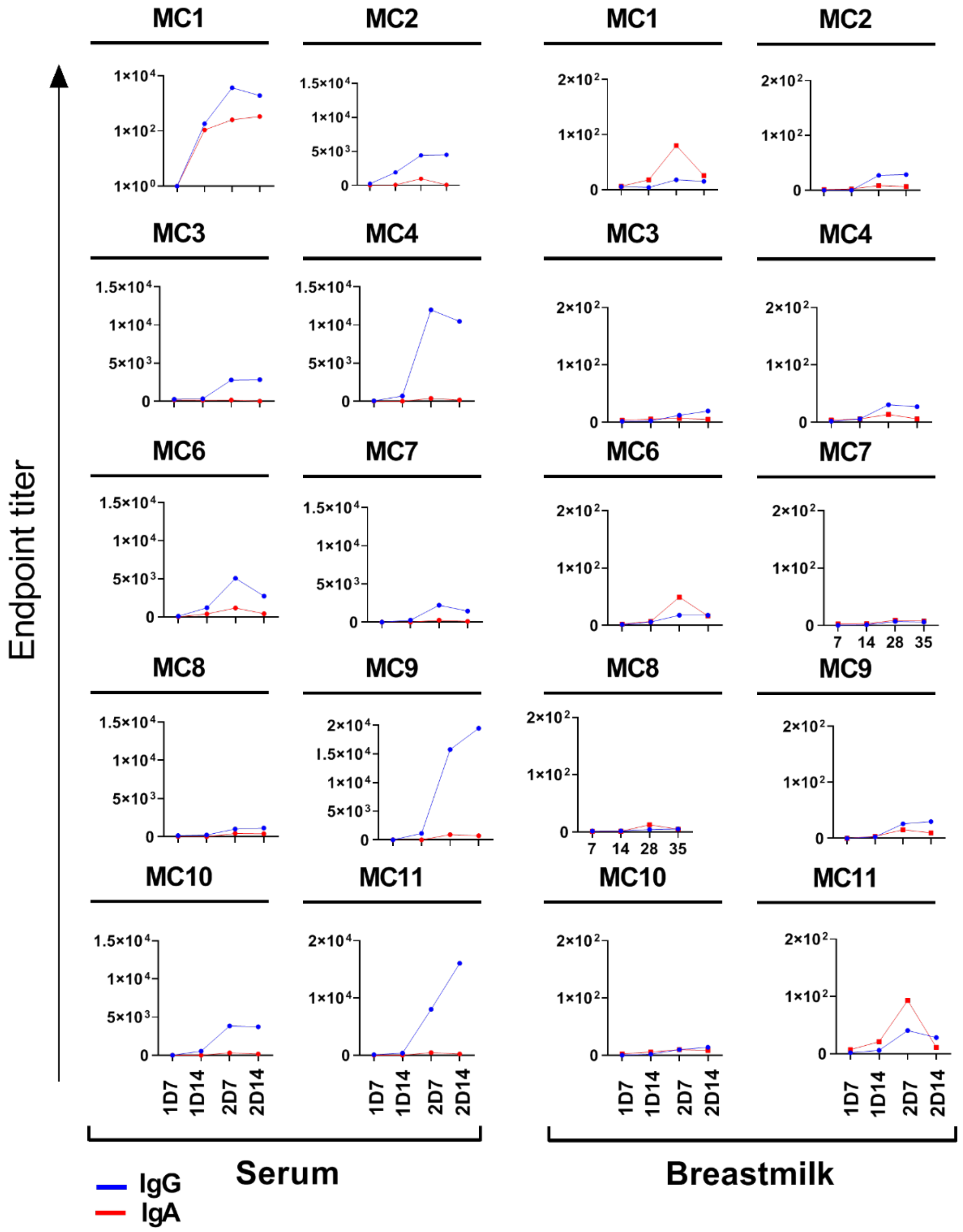


**Supplementary Fig. 2 | Temporal dynamics of spike and RBD specific antibody responses.** Endpoint titers were calculated and plotted by time points following the first and second vaccine doses for each participant. Breastmilk and serum samples were tested against SARS-CoV-2 spike (**A**) and RBD (**B**) proteins. Y-axis units are endpoint titers on a linear scale.

### Supplementary Fig.3

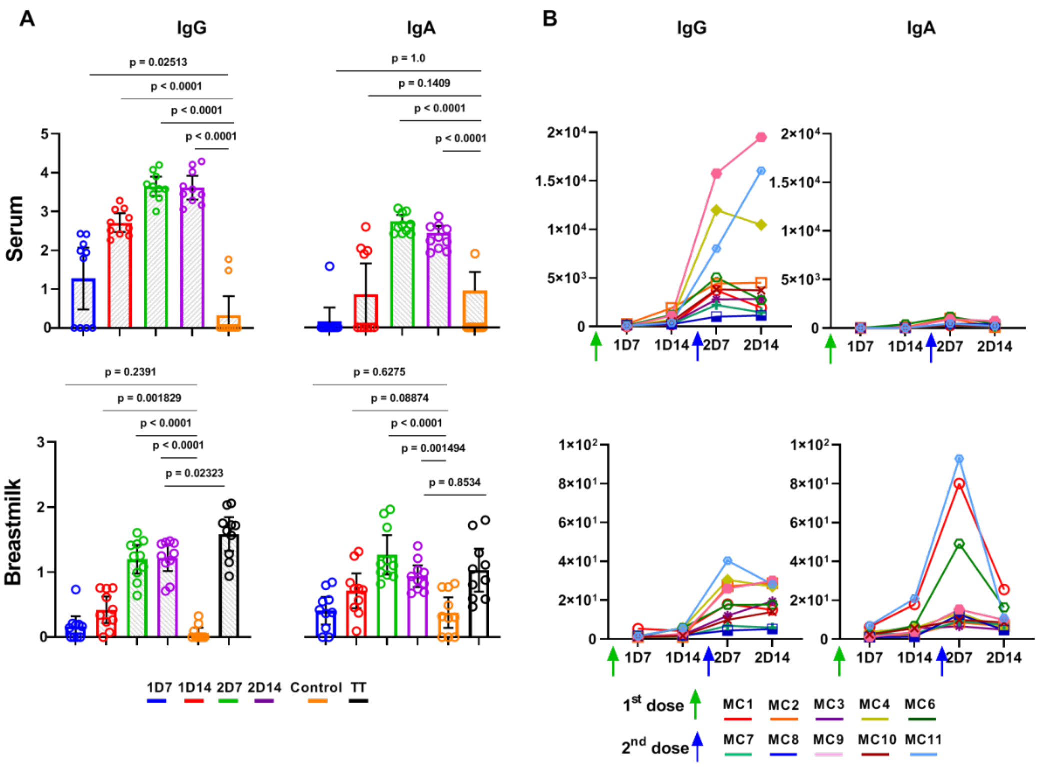


**Supplementary Fig. 3 | Endpoint titers of RBD-specific antibodies in breastmilk and serum (n=10).** (**A**) Comparison between the levels of vaccine-specific IgG and IgA antibodies as determined by ELISA endpoint titers. Endpoint titers were interpolated by applying a four parameter logistic curve (4PL) on reciprocal dilution series for all serum and lactoserum samples, at four time points (by color), for IgG and IgA. P values were determined with an unpaired, two-sided Mann-Whitney U-test after applying Bonferroni correction; $P < 0.0083$ was considered statistically significant. Results are presented as geometric means and 95% confidence intervals. Y-axis units are endpoint titers on a logarithmic scale. Control serum (n=10) and lactoserum (n=10) samples were obtained prior to the COVID-19 pandemic. (**B**) Endpoint titers per participant. Each colored line represents the titers for each participant. Green and blue arrows indicate the time points of administration of the first (t=0) and second (t=21) doses of the mRNA vaccine, respectively. Y-axis units are endpoint titers on a linear scale.

**Supplementary Fig.4**

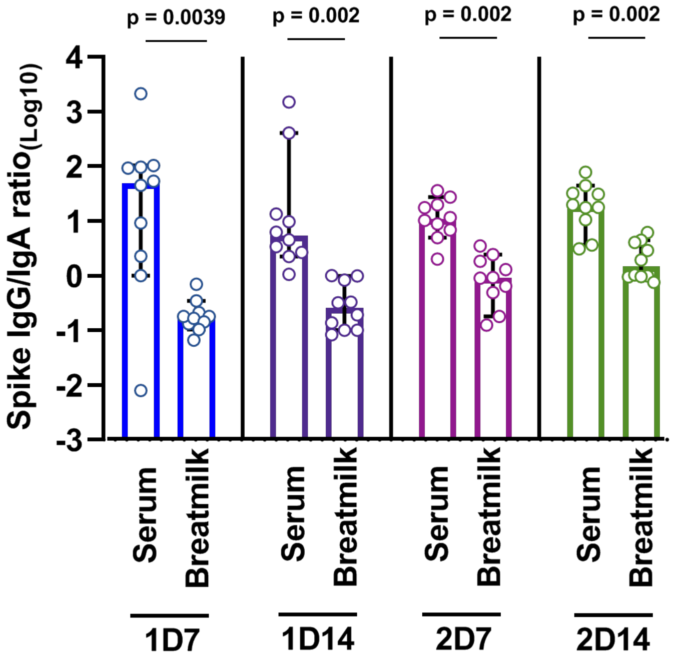


**Supplementary Fig. 4 | Spike-specific IgG/IgA titer ratio in breastmilk and serum at 4 time points following first and second doses of mRNA vaccine.** Comparisons between IgG/IgA ratio in serum vs. breastmilk were made using the two-sidesd Wilcoxon signed-rank test; $P < 0.05$ was considered statistically significant.

**Supplementary Fig.5**

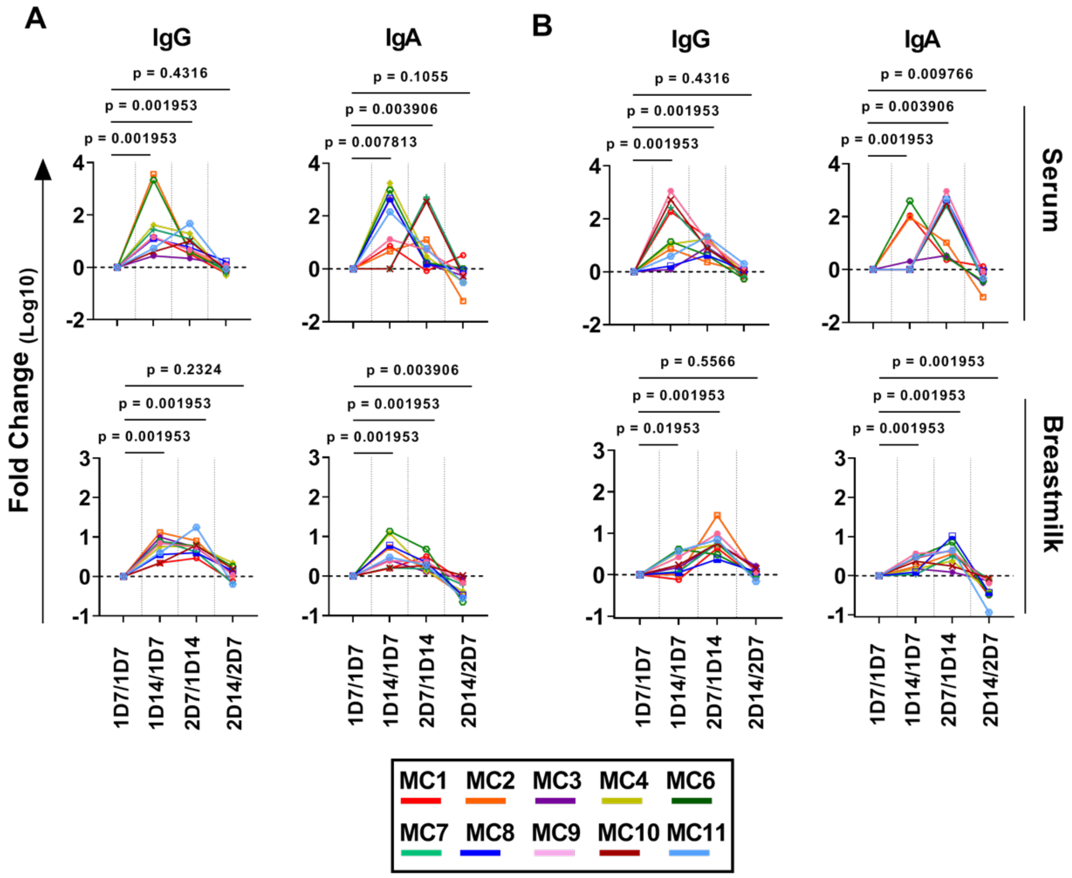


**Supplementary Fig. 5 | Fold-change of the endpoint vaccine-specific antibody titers.** Line graphs showing the endpoint titer fold-change in comparison to the proceeding time point for spike (**A**) and RBD (**B**) proteins. Comparisons between the fold-change in vaccine-specific IgG and IgA antibodies were made using the two-sidesd Wilcoxon signed-rank test; $P < 0.05$ was considered statistically significant. Y-axis units are fold-change in logarithmic scale.

**Supplementary Fig. 6**

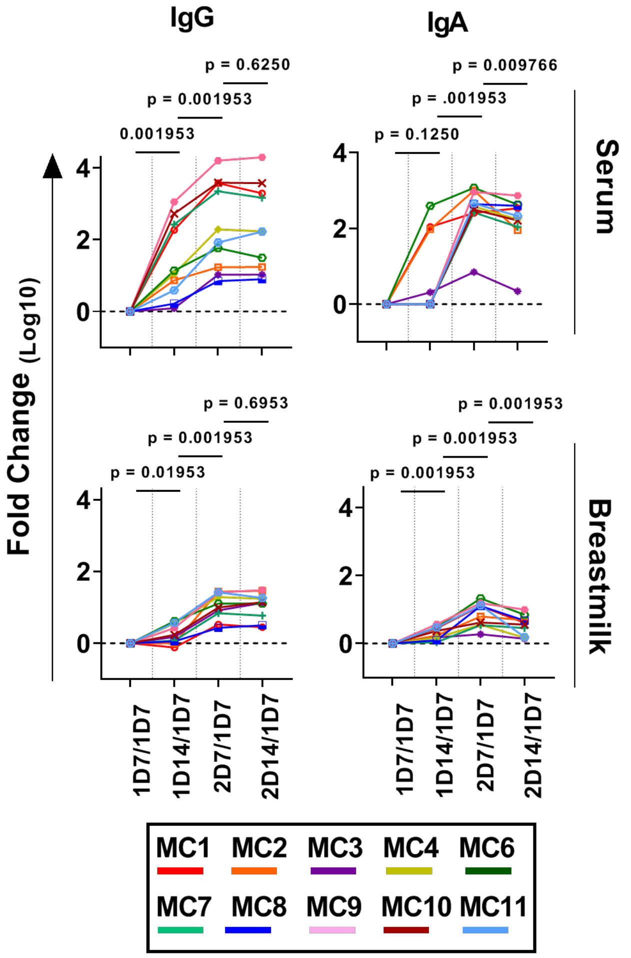


**Supplementary Fig. 6 | The dynamics of the RBD-specific antibody response.** Fold-change in antibody titers compared to the first time point (1D7) are plotted by patient. Comparisons between the fold-change in RBD-specific IgG and IgA antibodies were made using the two-sided Wilcoxon signed-rank test; $P < 0.05$ was considered statistically significant. Y-axis units are fold-change on a logarithmic scale.

**Supplementary Fig. 7**

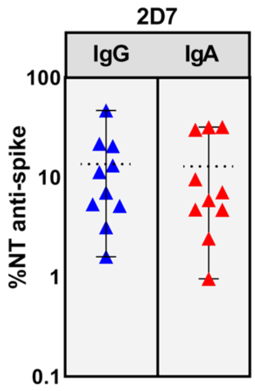


**Supplementary Fig. 7 | Fraction of spike-specific neutralizing antibodies in breastmilk.**

The percent of neutralizing (NT) antibodies out of the total spike-specific antibodies at the 2D7 time point as determined by a competitive-blocking assay with ACE2 as the competitor. Dashed black line indicates the mean fraction of neutralizing antibodies.
